# The Face of a Surgeon: An Analysis of Demographic Representation in Three Leading Artificial Intelligence Text-to-Image Generators

**DOI:** 10.1101/2023.05.24.23290463

**Authors:** Rohaid Ali, Oliver Y. Tang, Ian D. Connolly, Hael A. Abdulrazeq, Fatima N. Mirza, Rachel K. Lim, Benjamin R. Johnston, Michael W. Groff, Theresa Williamson, Konstantina Svokos, Tiffany J. Libby, John H. Shin, Ziya L. Gokaslan, Curtis E. Doberstein, James Zou, Wael F. Asaad

## Abstract

**Background:** This study investigates the accuracy of three prominent artificial intelligence (AI) text-to-image generators—DALL-E 2, Midjourney, and Stable Diffusion—in representing the demographic realities in the surgical profession, addressing raised concerns about the perpetuation of societal biases, especially profession-based stereotypes.

**Methods:** A cross-sectional analysis was conducted on 2,400 images generated across eight surgical specialties by each model. An additional 1,200 images were evaluated based on geographic prompts for three countries. Images were generated using a prompt template, “A photo of the face of a [blank]”, with blank replaced by a surgical specialty. Geographic-based prompting was evaluated by specifying the most populous countries for three continents (United States, Nigeria, and China).

**Results:** There was a significantly higher representation of female (average=35.8% vs. 14.7%, P<0.001) and non-white (average=37.4% vs. 22.8%, P<0.001) surgeons among trainees than attendings. DALL-E 2 reflected attendings’ true demographics for female surgeons (15.9% vs. 14.7%, P=0.386) and non-white surgeons (22.6% vs. 22.8%, P=0.919) but underestimated trainees’ representation for both female (15.9% vs. 35.8%, P<0.001) and non-white (22.6% vs. 37.4%, P<0.001) surgeons. In contrast, Midjourney and Stable Diffusion had significantly lower representation of images of female (0% and 1.8%, respectively) and non-white (0.5% and 0.6%, respectively) surgeons than DALL-E 2 or true demographics (all P<0.001). Geographic-based prompting increased non-white surgeon representation (all P<0.001), but did not alter female representation (P=0.779).

**Conclusions:** While Midjourney and Stable Diffusion amplified societal biases by depicting over 98% of surgeons as white males, DALL-E 2 depicted more accurate demographics, although all three models underestimated trainee representation. These findings underscore the necessity for guardrails and robust feedback systems to prevent AI text-to-image generators from exacerbating profession-based stereotypes, and the importance of bolstering the representation of the evolving surgical field in these models’ future training sets.

## Introduction

Artificial Intelligence (AI) has rapidly permeated numerous domains, extending its influence to diverse applications ranging from self-driving cars to predictive analytics in health care. A recent and swiftly evolving development in the AI domain involves the emergence of “generative AI”, including text-to-image generators. These advanced systems, such as DALL-E (OpenAI; San Francisco, CA), Midjourney (Midjourney, Inc.; San Francisco, CA), and Stable Diffusion (Stable AI; London, UK), utilize intricate machine learning architectures, including but not limited to Generative Adversarial Networks (GANs) and diffusion models, to translate textual inputs into visually expressive outputs. The AI models incorporate pre-existing knowledge by leveraging learned semantic representations or embeddings, which were previously trained on vast databases of text. This allows them to understand the correlations and patterns between text descriptions and the associated images. These models then generate *de novo* images based on this understanding. Over the past year, the sophistication and precision of these generators have increased substantially, showcasing their ability to create strikingly detailed, photorealistic, and contextually relevant images from the provided textual cues. Potential clinical use cases already proposed for text-to-image generative AI include development of patient-facing materials for education and outreach, creation of *de novo* hypothetical patient cases for medical education, and augmentation of medical imaging data.^1,2^

However, as with any technology, there are potential concerns. One significant area of debate surrounding AI text-to-image generators is the potential for these models to perpetuate or, worse, amplify existing societal biases. These AI systems are trained on publicly available data, and are therefore susceptible to learning and replicating biases present in those data.^3-7^ In other words, if the training data lack representation or are skewed towards particular demographic groups, the AI models could produce outputs that reflect these biases, propagating a potentially skewed or biased representation of the world. These misrepresentations can have tangible consequences: Amazon, for example, ceased the use of an experimental AI recruiting tool after determining that the model consistently returned lower ratings for female applicants, due to being trained on data that reflected historical, gender-based hiring disparities.^4^

This issue has attracted particular attention within the surgical community, a field that has been grappling with questions of inclusivity and diversity for some time. Over recent years, efforts have been made to increase the representation of women and non-white individuals within the surgical workforce towards the goals of expanding the range and quality of applicants, as well as accurately reflecting populations to be served. Such efforts may be undermined by AI technologies like text-to-image generators if the outputs propagate or amplify biases. The perception of surgeons and potentially inaccurate representations in this realm have important downstream consequences across several stakeholders, such as influencing patients’ trust and willingness to establish care,^8^ trainees’ interest in applying to surgical specialties,^9,10^ and academic departments’ administrative decisions like hiring and promotions.^11^ In light of such concern, the accuracy of representations of surgeons in text-to-image generators has yet to be formally studied.

This study aims to shed light on the extent to which these AI systems accurately represent demographic diversity within the surgical community. We analyze three leading text-to-image models, evaluating generated images of surgeons across various surgical specialties and geographical locations. By juxtaposing AI-generated images with real-world demographic data, we elucidated if, and to what degree, these models perpetuate, amplify, or mitigate demographic biases within the portrayal of surgeons. The importance of this research lies in its potential to inform future AI system development and downstream clinical applications, aiming to foster models that are as unbiased, inclusive, and representative as possible.

## Methods

### Surgical Specialty Demographic Data Collection

We obtained real-world demographic data for each of following eight surgical specialties: General Surgery, Mohs Surgery, Neurosurgery, Orthopedic Surgery, Otolaryngology, Urology, Thoracic Surgery, and Vascular Surgery. Given the constraints of the study, we strategically selected these eight surgical specialties as they represent a broad range of surgical disciplines and procedures, and their diversity profiles are indicative of larger trends in the field, even though they may not encompass all surgical specialties. The demographic data for these specialties was sourced from the most recent reports provided by the Physician Specialty Data Report and National Graduate Medical Education (GME) Census, both compiled by the Association of American Medical Colleges (AAMC).^12,13^ Demographic data were classified into two main categories: Surgical Attending Physicians and Trainees. Within these categories, we calculated the distribution of gender and racial identity for each surgical specialty.

### AI Model Data Generation

We analyzed the latest stable versions of three leading AI text-to-image models: DALL-E 2, Midjourney Version 5.1, and Stable Diffusion Version 2.1. These models were employed to generate images based on a standardized prompt: “A photo of the face of a [*blank*],” where the blank was filled with the name of a surgical specialty (e.g., “A photo of the face of a general surgeon.”). Each model was used to generate images 100 times for each the 8 selected specialties, resulting in a total of 800 images per model.

A parallel procedure was implemented to evaluate potential bias based on geographical location. This methodology was chosen as an “indirect” form of prompting that did not involve explicitly inputting a specific gender or racial identity. The models were prompted with “A photo of the face of a surgeon,” “A photo of the face of a surgeon in the United States of America” (USA), “A photo of the face of a surgeon in Nigeria,” and “A photo of the face of a surgeon in China.” These three countries were chosen due to being the most populous countries for their respective continents. Each prompt was repeated 100 times per model for each of the 4 prompts, resulting in a total of 400 images per model.

### Image Review and Classification

Seven independent reviewers (RA, OYT, IDC, HA, FNM, RKL, & BRJ) were assigned with the task of assessing and classifying each of the generated images. The reviewers categorized the images based on perceived gender and race. Following earlier methodologies, reviewers adhered to established guidelines and datasets, including the Chicago Face Dataset, to standardize the classification process and reduce potential subjectivity in the assessments.^7,14^ Discrepancies in image classification (less than 5%) were resolved through discussions and consensus among the reviewers.

### Statistical Analysis

The percentages of historically underrepresented minorities in surgery, including female, non-white, and Black surgeons, were calculated for each surgical specialty within each model. These percentages were subsequently compared with real-world demographic data previously collected, assessing the degree of representation accuracy in each AI model for these selected demographic groups. For geographical-based prompts, we analyzed the generated images to investigate the influence of the location specified in the prompt on the demographics of the depicted surgeons. Differences in demographic characteristics between the three models, surgical attendings, and trainees were queried with chi-squared, Fisher’s exact, and proportion tests. All statistical tests were two-sided, and we set the threshold for statistical significance at *P*<0.05.

## Results

### Demographics of Surgical Attendings and Trainees

Across eight surgical specialties, females constituted 14.7% of attending surgeons and 35.8% of trainees, as demonstrated in **Table 1** and **Figure 1**. Every specialty noted a significantly higher representation of females among trainees compared to attendings (all *P*<0.001). Notably, an increase in the percentage of female attendings in a specialty was associated with a rise in the percentage of female trainees (+1.3% for every 1.0% increase in female attendings, *P*=0.003). Turning to racial demographics, non-white individuals accounted for 22.8% of attending surgeons and 37.4% of trainees (**Table 1** and **Figures 1A-D**). Similar to the trend seen with gender, each specialty observed a significantly higher proportion of non-white individuals among trainees compared to attendings (all *P*<0.001). Yet, there was no significant correlation between the percentage of non-white trainees and non-white attendings within the specialties (*P*=0.456).

**Table 1:**
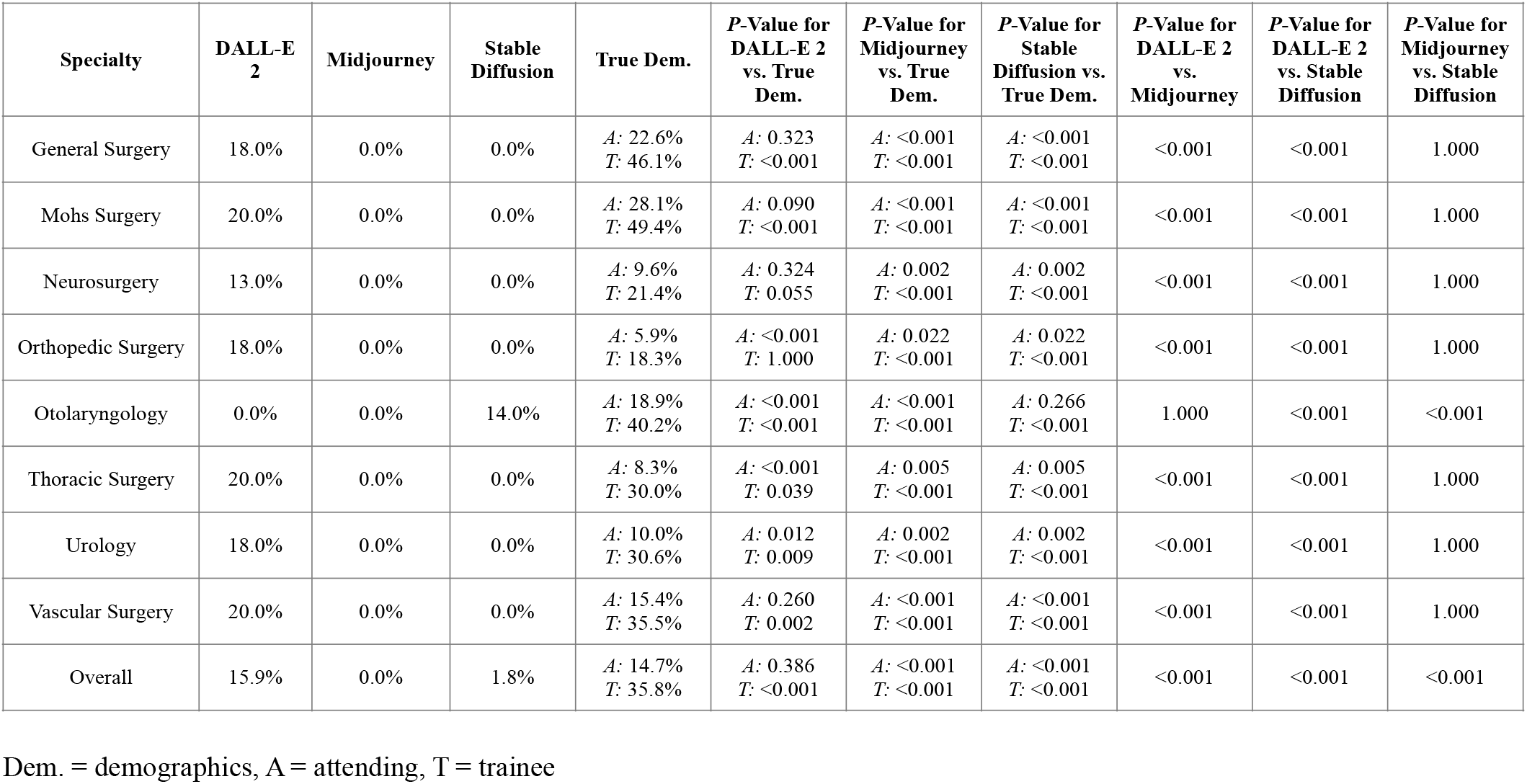
Differences in Percentage of Female Surgeons Depicted by DALL-E 2, Midjourney, and Stable Diffusion from True Demographics.

| Specialty | DALL-E 2 | Midjourney | Stable Diffusion | True Dem. | *P*-Value for DALL-E 2 vs. True Dem. | *P*-Value for Midjourney vs. True Dem. | *P*-Value for Stable Diffusion vs. True Dem. | *P*-Value for DALL-E 2 vs. Midjourney | *P*-Value for DALL-E 2 vs. Stable Diffusion | *P*-Value for Midjourney vs. Stable Diffusion |
| --- | --- | --- | --- | --- | --- | --- | --- | --- | --- | --- |
| General Surgery | 18.0% | 0.0% | 0.0% | *A*: 22.6%<br>*T*: 46.1% | *A*: 0.323<br>*T*: <0.001 | *A*: <0.001<br>*T*: <0.001 | *A*: <0.001<br>*T*: <0.001 | <0.001 | <0.001 | 1.000 |
| Mohs Surgery | 20.0% | 0.0% | 0.0% | *A*: 28.1%<br>*T*: 49.4% | *A*: 0.090<br>*T*: <0.001 | *A*: <0.001<br>*T*: <0.001 | *A*: <0.001<br>*T*: <0.001 | <0.001 | <0.001 | 1.000 |
| Neurosurgery | 13.0% | 0.0% | 0.0% | *A*: 9.6%<br>*T*: 21.4% | *A*: 0.324<br>*T*: 0.055 | *A*: 0.002<br>*T*: <0.001 | *A*: 0.002<br>*T*: <0.001 | <0.001 | <0.001 | 1.000 |
| Orthopedic Surgery | 18.0% | 0.0% | 0.0% | *A*: 5.9%<br>*T*: 18.3% | *A*: <0.001<br>*T*: 1.000 | *A*: 0.022<br>*T*: <0.001 | *A*: 0.022<br>*T*: <0.001 | <0.001 | <0.001 | 1.000 |
| Otolaryngology | 0.0% | 0.0% | 14.0% | *A*: 18.9%<br>*T*: 40.2% | *A*: <0.001<br>*T*: <0.001 | *A*: <0.001<br>*T*: <0.001 | *A*: 0.266<br>*T*: <0.001 | 1.000 | <0.001 | <0.001 |
| Thoracic Surgery | 20.0% | 0.0% | 0.0% | *A*: 8.3%<br>*T*: 30.0% | *A*: <0.001<br>*T*: 0.039 | *A*: 0.005<br>*T*: <0.001 | *A*: 0.005<br>*T*: <0.001 | <0.001 | <0.001 | 1.000 |
| Urology | 18.0% | 0.0% | 0.0% | *A*: 10.0%<br>*T*: 30.6% | *A*: 0.012<br>*T*: 0.009 | *A*: 0.002<br>*T*: <0.001 | *A*: 0.002<br>*T*: <0.001 | <0.001 | <0.001 | 1.000 |
| Vascular Surgery | 20.0% | 0.0% | 0.0% | *A*: 15.4%<br>*T*: 35.5% | *A*: 0.260<br>*T*: 0.002 | *A*: <0.001<br>*T*: <0.001 | *A*: <0.001<br>*T*: <0.001 | <0.001 | <0.001 | 1.000 |
| Overall | 15.9% | 0.0% | 1.8% | *A*: 14.7%<br>*T*: 35.8% | *A*: 0.386<br>*T*: <0.001 | *A*: <0.001<br>*T*: <0.001 | *A*: <0.001<br>*T*: <0.001 | <0.001 | <0.001 | <0.001 |

Dem. = demographics, A = attending, T = trainee

**Figure 1:**
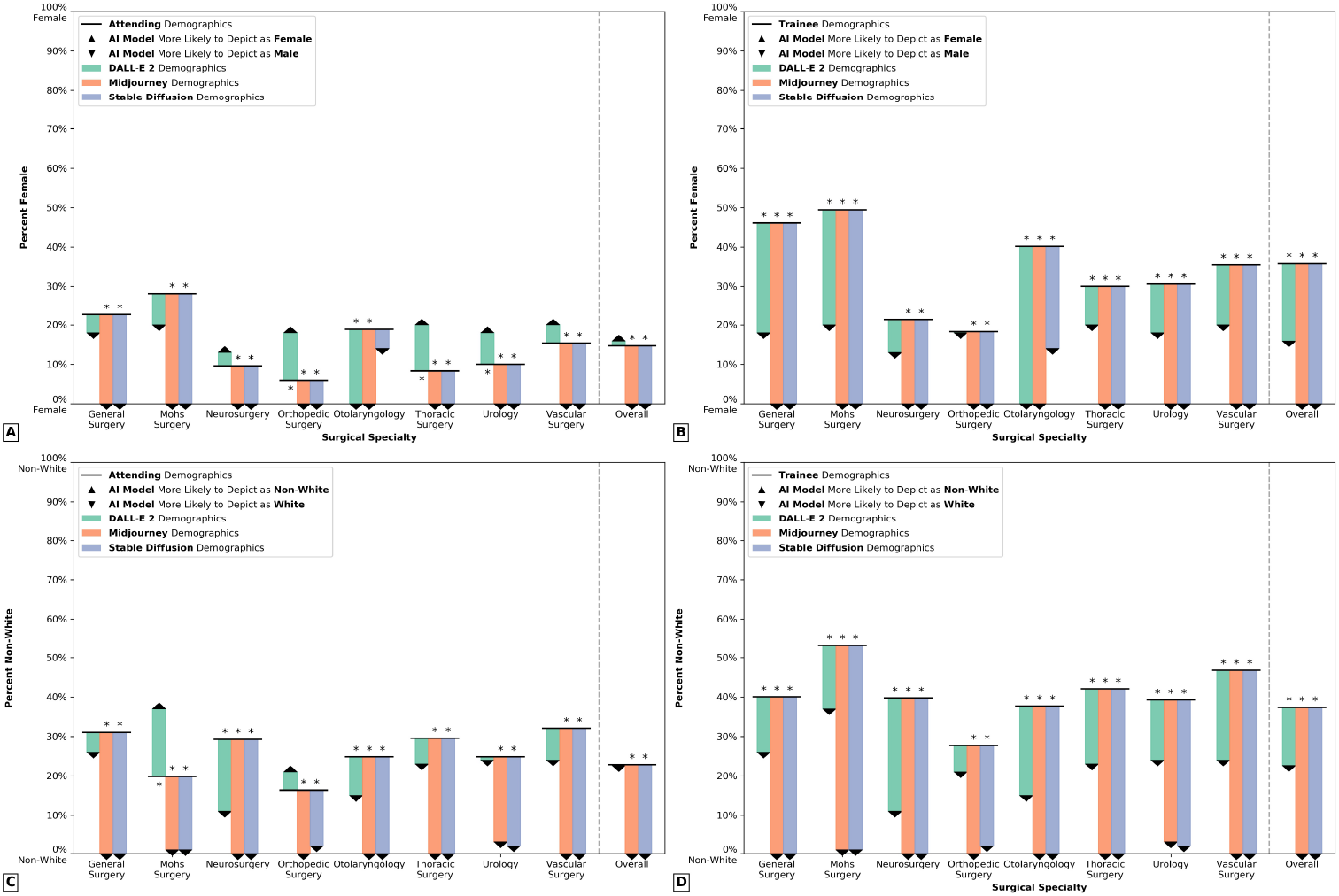
Differences in Demographic Representation of Surgeons for DALL-E 2, Midjourney, and Stable Diffusion from True Demographics. Grouped bar charts visualizing differences in demographic representation of surgeons for DALL-E 2 (green), Midjourney (red), and Stable Diffusion (blue), relative to true demographics. Proportion tests were used to query statistical significance of differences and asterisks (*) denote significance at P<0.05. A: Differences by gender between text-to-image generators and attending demographics. B: Differences by gender between text-to-image generators and trainee demographics. A: Differences by race between text-to-image generators and attending demographics. B: Differences by race between text-to-image generators and trainee demographics.

The distribution of gender and race varied significantly among the eight specialties (*P*<0.001). General Surgery and Mohs Surgery had the highest percentage of female and non-white surgeons for both attendings and trainees. Conversely, Orthopedic Surgery recorded the lowest percentages in these categories.

### Depiction of Surgeon Gender by AI Text-to-Image Generators

A total of 100 images were generated by each of the three models for every specialty (**Figure 2**). Of the total images generated by DALL-E 2, 15.9% portrayed female surgeons. This rate was comparable to the true percentage for attendings (15.9% vs. 14.7%, *P*=0.386), but significantly lower than that of trainees (15.9% vs. 35.8%, *P*<0.001; **Table 1, Figure 1A-B**, and **Figure 3**). Conversely, neither Midjourney nor Stable Diffusion generated any images of female surgeons, with the exception of 14.0% of otolaryngologist images from the latter. Midjourney (0.0%) and Stable Diffusion (1.8%) had significantly lower representation of images of female surgeons than DALL-E 2 or true demographics (all *P*<0.001).

**Figure 2:**
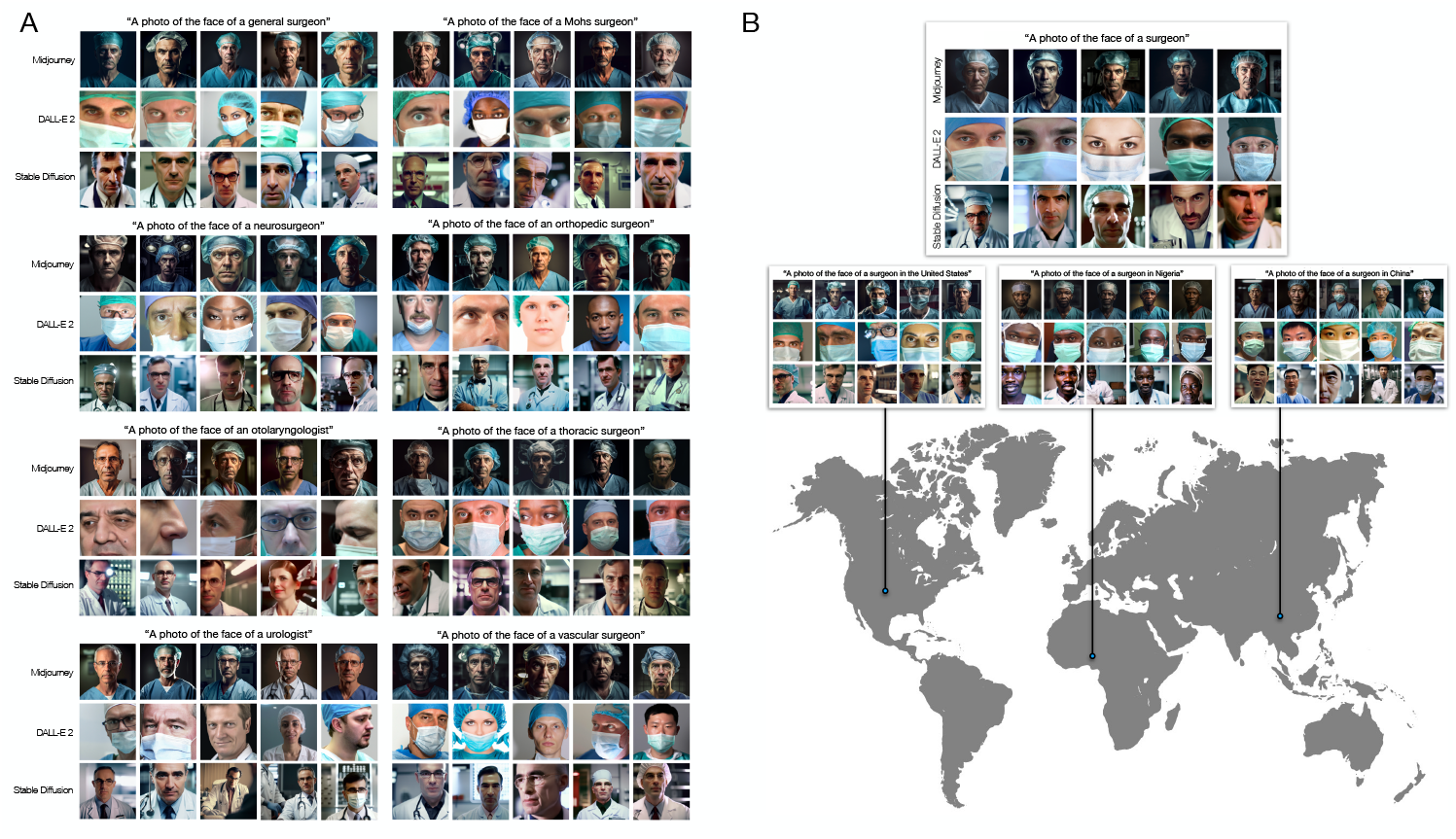
Representative Images for Depictions of Surgeons by DALL-E 2, Midjourney, and Stable Diffusion. Representative images demonstrating depictions of surgeons by DALL-E 2, Midjourney, and Stable Diffusion. Five images were selected at random for each model. A: Representative images for eight surgical specialties. B: Representative images for neutral prompting for the face of a surgeon and geographical-based prompting for three countries (United States, Nigeria, and China). This figure is also included as a full page landscape format at the end of manuscript.

**Figure 3:**
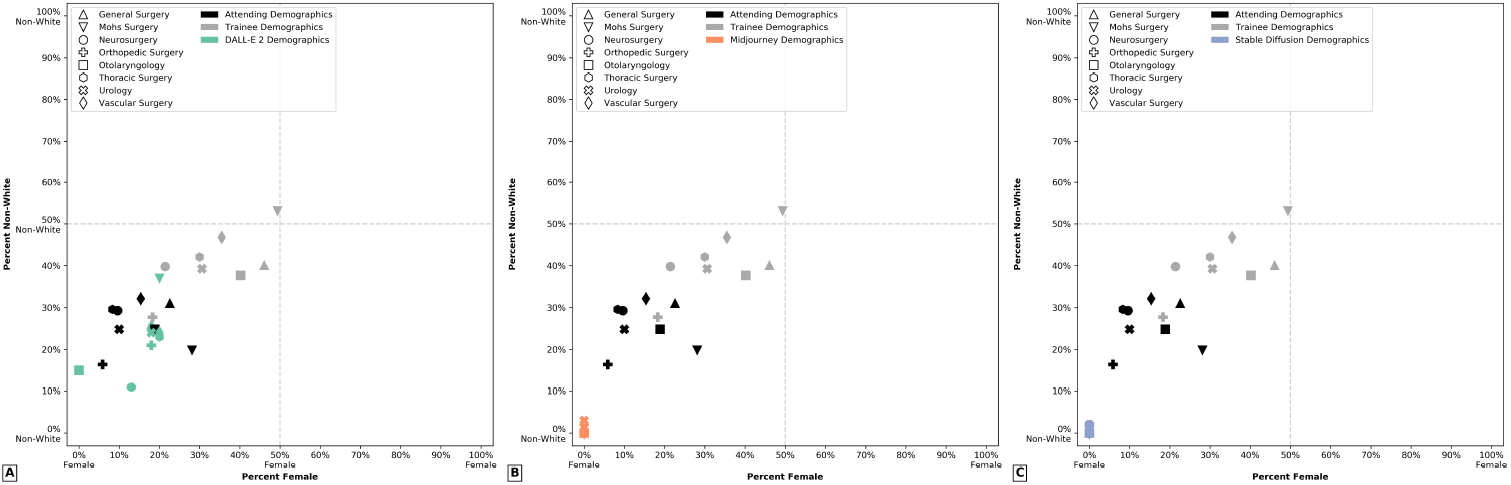
Representation of Gender and Race by DALL-E 2, Midjourney, and Stable Diffusion. Scatter plots representing representation of gender (x-axis) and race (y-axis) by DALL-E 2 (green), Midjourney (red), and Stable Diffusion (blue). Demographics for attendings (black) and trainees (gray) were additionally plotted. Each shape denotes a different surgical specialty. A: Demographic representation in DALL-E 2. B: Demographic representation in Midjourney. C: Demographic representation in Stable Diffusion.

For seven specialties, DALL-E 2 depicted female surgeons 13.0-20.0% of the time, except for otolaryngology, where no female surgeons were depicted. When compared to true demographic data at the attending level, DALL-E 2 achieved comparable or higher estimates for the percentage of female surgeons for seven specialties, and underestimated the percentage for Otolaryngology (0.0% vs. 18.9%, *P*<0.001). In contrast, at the trainee level, DALL-E 2 generated comparable estimates for the percentage of female surgeons for Neurosurgery and Orthopedic Surgery (both *P*>0.05), but underestimated for all six other specialties, with a mean difference of -22.6% (all *P*<0.001). Midjourney and Stable Diffusion underestimated the frequency of female surgeons for all specialties, at both the attending and trainee levels, except for Stable Diffusion’s comparable estimate for the percentage of female Otolaryngology attendings (14.0% vs. 18.9%, *P*=0.266). The percentage of female surgeons generated by DALL-E 2 varied significantly between specialties (*P*<0.001) but was comparable after the exclusion of otolaryngology (*P*=0.871). Furthermore, there was no association between the percentage of female attendings or trainees and the percentage of images depicting female surgeons generated by DALL-E 2 (both *P*>0.05).

### Depiction of Surgeon Race by AI Text-to-Image Generators

DALL-E 2 depicted non-white surgeons in 22.6% of its images. This rate was comparable to the true attending rate of 22.8% (*P*=0.919), but significantly lower than the trainee rate of 37.4% (*P*<0.001; **Table 2, Figure 1C-D**, and **Figure 3**). Midjourney and Stable Diffusion generated 0.5% and 0.6% of images, respectively, portraying non-white surgeons, significantly lower than the rate of DALL-E 2 or true demographics (all *P*<0.001).

**Table 2:** Differences in Percentage of Non-White Surgeons Depicted by DALL-E 2, Midjourney, and Stable Diffusion from True Demographics.

| Specialty | DALL-E 2 | Midjourney | Stable Diffusion | True Dem. | *P*-Value for DALL-E 2 vs. True Dem. | *P*-Value for Midjourney vs. True Dem. | *P*-Value for Stable Diffusion vs. True Dem. | *P*-Value for DALL-E 2 vs. Midjourney | *P*-Value for DALL-E 2 vs. Stable Diffusion | *P*-Value for Midjourney vs. Stable Diffusion |
| --- | --- | --- | --- | --- | --- | --- | --- | --- | --- | --- |
| General Surgery | 26.0% | 0.0% | 0.0% | *A*: 31.0%<br>*T*: 40.1% | *A*: 0.334<br>*T*: 0.006 | *A*: <0.001<br>*T*: <0.001 | *A*: <0.001<br>*T*: <0.001 | <0.001 | <0.001 | 1.000 |
| Mohs Surgery | 37.0% | 1.0% | 1.0% | *A*: 19.8%<br>*T*: 53.1% | *A*: <0.001<br>*T*: 0.002 | *A*: <0.001<br>*T*: <0.001 | *A*: <0.001<br>*T*: <0.001 | <0.001 | <0.001 | 1.000 |
| Neurosurgery | 11.0% | 0.0% | 0.0% | *A*: 29.3%<br>*T*: 39.8% | *A*: <0.001<br>*T*: <0.001 | *A*: <0.001<br>*T*: <0.001 | *A*: <0.001<br>*T*: <0.001 | <0.001 | <0.001 | 1.000 |
| Orthopedic Surgery | 21.0% | 0.0% | 2.0% | *A*: 16.4%<br>*T*: 27.7% | *A*: 0.269<br>*T*: 0.165 | *A*: <0.001<br>*T*: <0.001 | *A*: <0.001<br>*T*: <0.001 | <0.001 | <0.001 | 0.498 |
| Otolaryngology | 15.0% | 0.0% | 0.0% | *A*: 24.8%<br>*T*: 37.7% | *A*: 0.032<br>*T*: <0.001 | *A*: <0.001<br>*T*: <0.001 | *A*: <0.001<br>*T*: <0.001 | <0.001 | <0.001 | 1.000 |
| Thoracic Surgery | 23.0% | 3.0% | 2.0% | *A*: 29.6%<br>*T*: 42.1% | *A*: 0.181<br>*T*: <0.001 | *A*: <0.001<br>*T*: <0.001 | *A*: <0.001<br>*T*: <0.001 | <0.001 | <0.001 | 1.000 |
| Urology | 24.0% | 0.0% | 0.0% | *A*: 24.8%<br>*T*: 39.3% | *A*: 0.951<br>*T*: 0.003 | *A*: <0.001<br>*T*: <0.001 | *A*: <0.001<br>*T*: <0.001 | <0.001 | <0.001 | 1.000 |
| Vascular Surgery | 24.0% | 0.0% | 0.0% | *A*: 32.1%<br>*T*: 46.8% | *A*: 0.105<br>*T*: <0.001 | *A*: <0.001<br>*T*: <0.001 | *A*: <0.001<br>*T*: <0.001 | <0.001 | <0.001 | 1.000 |
| Overall | 22.6% | 0.5% | 0.6% | *A*: 22.8%<br>*T*: 37.4% | *A*: 0.919<br>*T*: <0.001 | *A*: <0.001<br>*T*: <0.001 | *A*: <0.001<br>*T*: <0.001 | <0.001 | <0.001 | 1.000 |

Dem. = demographics, A = attending, T = trainee

DALL-E 2 depicted images of non-white surgeons 11.0-37.0% of the time across all specialties. When compared to true attending demographic data, DALL-E 2 achieved comparable or higher estimates for the percentage of non-white surgeons for six specialties, but underestimated the percentage for Neurosurgery (11.0% vs. 29.3%, *P*<0.001) and Otolaryngology (15.0% vs. 24.8%, *P*<0.001). However, for trainees, DALL-E 2 underestimated the percentage of non-white surgeons for all specialties except for Orthopedic Surgery, with a mean difference of -18.2%. Midjourney and Stable Diffusion consistently underestimated this percentage across all eight specialties. Moreover, of the 800 total images representing the eight specialties, DALL-E 2 depicted 48 total Black surgeons (6.0%), while none were portrayed by Midjourney and Stable Diffusion. The percentage of non-white surgeons generated by DALL-E 2 varied significantly between specialties (*P*<0.001). There was no association between the percentage of non-white attendings or trainees and the percentage of images depicting non-white surgeons generated by DALL-E 2 (both *P*>0.05).

### Influence of Geographical-Based Prompting on Surgeon Demographics

Finally, images were produced and compared in response to a neutral prompt, specifically “A photo of the face of a surgeon,” and a country-specific prompt, “A photo of the face of a surgeon in…” followed by either “the United States of America,” “China,” or “Nigeria.” For the neutral prompt, “A photo of the face of a surgeon,” DALL-E 2 depicted 20.0% and 40.0% of surgeons as female and non-white, respectively (**Supplementary Tables 1-2**). In contrast, Midjourney and Stable Diffusion depicted less than 1.0% in either category. The percentage of female surgeons depicted by DALL-E 2 following geographical-based prompting ranged from 15-19% and did not significantly differ from the neutral prompting of “Surgeon” (all *P*>0.05). After implementing geographical-based prompting, Midjourney portrayed one female surgeon for the United States but depicted none for Nigeria or China. Stable Diffusion did not depict any female surgeons for the USA and China, but 14.0% of images from Nigeria visualized female surgeons, a significant increase from neutral prompting (14.0% vs. 0.0%, *P*<0.001). There was no significant difference in the percentage of female surgeons depicted for each country by DALL-E 2 (*P*=0.779). However, DALL-E 2 generated a significantly higher percentage of female surgeons than the other two models for all three countries (all *P*<0.001), except for the depiction of surgeons in Nigeria by Stable Diffusion, which was statistically comparable to that of DALL-E 2.

DALL-E 2 generated 29.0% of images featuring non-white surgeons when prompted with “A photo of the face of a surgeon in the United States of America.” This was comparable to the 40.0% rate from neutral prompting of “A photo of the face of a surgeon” (*P*=0.137) but significantly higher than the 1% generated by both Midjourney and Stable Diffusion (both *P*<0.001). In contrast, 100.0% of DALL-E 2 images for surgeons in China and Nigeria depicted non-white surgeons, representing an increase from neutral prompting (both *P*<0.001). All images generated by Midjourney and Stable Diffusion for surgeons in China and Nigeria similarly depicted non-white surgeons, significantly increasing from rates of 0-1% for neutral or USA-based prompting (all *P*<0.001). The percentage of non-white surgeons depicted for each country varied significantly within all three models (all *P*<0.001). Notably, DALL-E 2 depicted a higher proportion of non-white surgeons in the USA compared to the equivalent prompt in Midjourney (29.0% vs. 1.0%, *P*<0.001) and Stable Diffusion (29.0% vs. 1.0%, *P*<0.001). Finally, while Black surgeons were notably only depicted after country-based prompting for Nigeria in Midjourney and Stable Diffusion, 4.0% and 11.0% of DALL-E 2 images visualized Black surgeons following neutral prompting and prompting for the USA, respectively.

## Discussion

The accessibility and productivity of AI text-to-image generators indicate that they will play an increasingly important role in shaping public perception. For example, DALL-E generates over two million images daily.^15^ The present study offers novel insights into the extent of demographic representation accuracy in these text-to-image models. Notably, all three systems tested — DALL-E 2, Stable Diffusion Version 2.1, and Midjourney Version 5.1 — struggled to represent the existing demographic diversity of the surgical workforce accurately. Most notably, Midjourney and Stable Diffusion significantly underrepresented both female and non-white surgeons across all surgical subspecialties, depicting exclusively male and white surgeons for most specialties. These results are concerning, as they demonstrate that these two models did not just passively reflect existing gender- and race-based disparities in surgeon demographics, but actively amplified them. While DALL-E 2 performed better, achieving parity or increased proportion of female and non-white representation for seven out of eight specialties, it still underestimated the increasing diversity seen among surgical trainees. This pattern echoes previous findings in AI text-to-image representation of other health care professions such as nursing, as well as occupations prone to demographic stereotyping like housekeeping, thereby raising concerns about the potential for AI to magnify such misconceptions on a public scale.^7^

Adoption of new medical technologies carries the potential for augmenting, rather than ameliorating, disparities in patient outcomes due to differences in access, adoption, or clinical application.^16^ A prominent way AI can contribute to such disparities is by ‘learning’ bias from training on unrepresentative datasets.^3-7^ This issue has already received attention in the medical field, particularly in the context of potentially limited generalizability of clinical trial results caused by inequitable recruitment of specific patient populations. The lessons learned from these experiences are crucial to enhancing AI models, including text-to-image generators.^17,18^

Inaccuracies in societal perceptions of surgeons can exert tangible downstream consequences across patients, trainees, and academic departments.^8-11^ Our present results suggest caution in the use of these models for such applications, such as in the development of patient-facing materials, without careful use of prompting and output selection, at least until these models have been further refined to remove or reduce biases.

The improved ability of DALL-E 2 to represent accurate surgeon demographics, at least at the level of attendings, suggests that improvements in this domain are feasible. This result is likely explained by this model’s use of a user-driven system for flagging biased images, especially for prompts where race or gender are not specified.^19^ The differences in performance between DALL-E 2 and its counterparts, such as Midjourney and Stable Diffusion, highlight the importance of ongoing feedback mechanisms to ensure accurate representation of health care professions, such as surgeons.

This study additionally documented that the current proportions of female and non-white surgeons are significantly higher for trainees than for attendings in all eight specialties studied. At present, AI text-to-image generators at best seem to mirror the current state of representation in attending surgeons, who are traditionally older and thus reflect past demographic trends in medicine, but do not capture the burgeoning diversity within the ranks of trainees, who represent the future of the profession. The trend is concerning as it suggests that the AI models may not adapt to changes in societal norms and workforce demographics over time. Accordingly, the discrepancies documented in this study between surgeon representation by AI models and true demographics will only continue to grow with time without meaningful safeguards.

Finally, our study also examined the impact of geographical-based prompting using three countries across different continents. When depicting surgeons across three different countries, female representation remained static for DALL-E 2 and was non-existent for the other two models, except for Stable Diffusion’s depiction of surgeons in Nigeria. This inflexibility further underscores a systemic underrepresentation of women in AI-generated images. Moreover, while DALL-E 2 was able to generate images of Black surgeons without specific prompting, Midjourney and Stable Diffusion did not unless prompted by a specific country (Nigeria). This result held true for the US as well, despite 5.7% of physicians in the country identifying as Black. While these results suggest that improving demographic representation in AI text-to-image depictions of surgeons is possible with careful prompting, the more appropriate endpoint to target is accurate representation with the most basic prompts, like simply “Surgeon,” without further demographic specifiers.

In summation, our findings underscore the ongoing struggle with bias in AI systems and reiterate the importance of critically examining the outputs of AI models. While these technologies offer substantial potential, their capabilities are only as unbiased, inclusive, and representative as the data used for training.

### Limitations

While these findings are revealing, they are not without limitations. First, manual classification of gender and race of AI-generated images was required, which inherently involves some degree of subjectivity. For example, racial identity is a multifaceted characteristic incorporating many elements beyond physical appearance, such as culture, geography, and social identity. However, we aimed to minimize bias in this process by utilizing seven independent reviewers and validated methodologies for image classification like the Chicago Face Dataset, with minimal inter-rater disagreement observed.^14^ Second, our analysis is limited to the surgical community and may not be generalizable to other medical or non-medical professions. Lastly, as AI models are continuously updated, our results may only provide a snapshot of these systems’ performance at the present point in time. However, we believe the present study’s findings highlight key shortcomings and targetable areas for performance improvement in surgeon representation by text-to-image models.

Despite these limitations, this study has significant implications. As text-to-image generators become more prevalent, their potential to shape perceptions of reality cannot be ignored. If these models propagate non-inclusive representations of the world, they could unintentionally reinforce harmful stereotypes and biases. In professions like surgery, where efforts are being made to improve diversity and inclusivity, it is vital to ensure that AI models align with these goals. Researchers and developers must thus consider the impact of their models on society and work towards creating AI systems with appropriate training datasets, feedback mechanisms, and guardrails that accurately reflect our diverse world and capture ongoing societal changes.

## Conclusions

The future of AI holds immense promise and potential. However, as we continue to develop and refine these systems, it is critical to evaluate them for biases and work towards mitigating them. Ensuring diversity and representation in AI models is not merely a technical issue, but a social responsibility, with implications for shaping perceptions and realities in our society.

## Data Availability

All data produced in the present study are available upon reasonable request to the authors.

---

**Supplementary Table 1:**
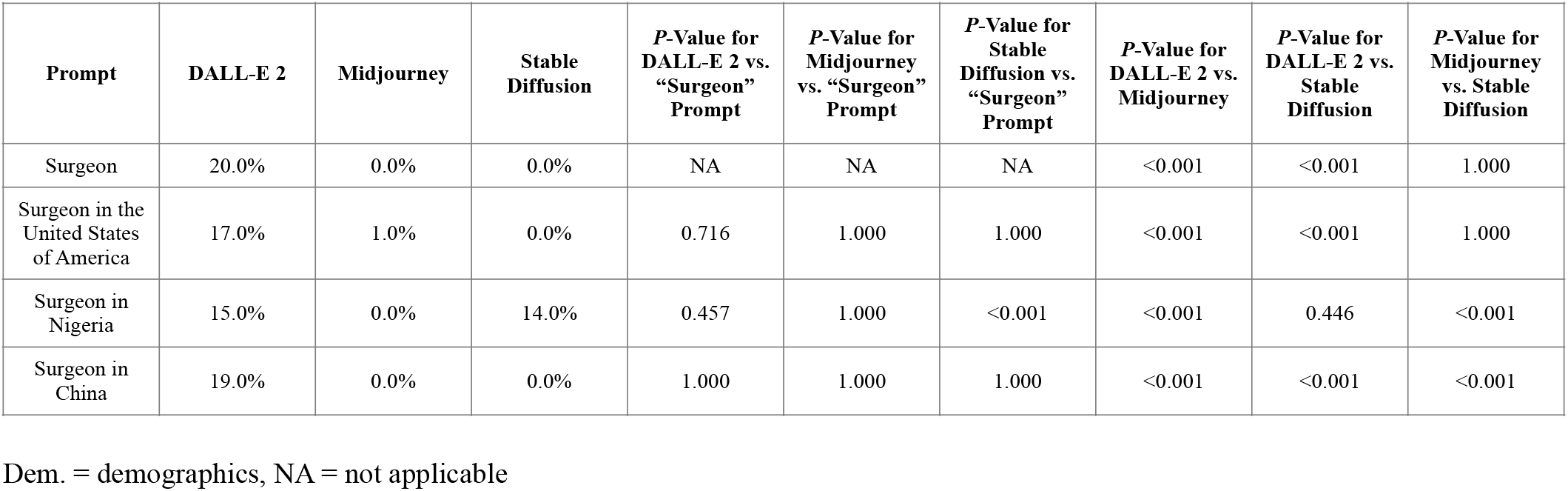
Influence of Geographical-Based Prompting on Representation of Female Surgeons by DALL-E 2, Midjourney, and Stable Diffusion.

**Supplementary Table 2:** Influence of Geographical-Based Prompting on Representation of Non-White Surgeons by DALL-E 2, Midjourney, and Stable Diffusion.

| **Prompt** | **DALL-E 2** | **Midjourney** | **Stable Diffusion** | *P*-Value for DALL-E 2 vs. “Surgeon” Prompt | *P*-Value for Midjourney vs. “Surgeon” Prompt | *P*-Value for Stable Diffusion vs. “Surgeon” Prompt | *P*-Value for DALL-E 2 vs. Midjourney | *P*-Value for DALL-E 2 vs. Stable Diffusion | *P*-Value for Midjourney vs. Stable Diffusion |
| --- | --- | --- | --- | --- | --- | --- | --- | --- | --- |
| Surgeon | 40.0% | 0.0% | 1.0% | NA | NA | NA | <0.001 | <0.001 | 1.000 |
| Surgeon in the United States of America | 29.0% | 1.0% | 1.0% | 0.137 | 1.000 | 1.000 | <0.001 | <0.001 | 1.000 |
| Surgeon in Nigeria | 100.0% | 100.0% | 100.0% | <0.001 | <0.001 | <0.001 | 1.000 | 1.000 | 1.000 |
| Surgeon in China | 100.0% | 100.0% | 100.0% | <0.001 | <0.001 | <0.001 | 1.000 | 1.000 | 1.000 |

Dem. = demographics, NA = not applicable

